# Advanced ECG heart age estimation applicable to both sinus and non-sinus rhythm associates with cardiovascular risk, cardiovascular morbidity, and survival

**DOI:** 10.1101/2024.03.12.24304123

**Authors:** Zaidon Al-Falahi, Todd T Schlegel, Israel Lamela-Palencia, Annie Li, Erik B Schelbert, Louise Niklasson, Maren Maanja, Thomas Lindow, Martin Ugander

**Affiliations:** Kolling Institute, Royal North Shore Hospital, and University of Sydney, Sydney, Australia; Department of Clinical Physiology, Karolinska University Hospital, and Karolinska Institutet, Stockholm, Sweden; Nicollier-Schlegel SARL, Trélex, Switzerland; Minneapolis Heart Institute East, United Hospital, Minnesota, USA; Clinical Physiology, Clinical Sciences, Lund University, Lund, Sweden

**Keywords:** ECG, accelerated ageing, advanced ECG analysis, machine learning, risk prediction

## Abstract

**Background:** An explainable advanced electrocardiography (A-ECG) heart age gap is the difference between A-ECG heart age and chronological age. This gap is an estimate of accelerated cardiovascular ageing expressed in years of healthy human aging, and can intuitively communicate cardiovascular risk to the general population. However, existing A-ECG heart age measures require discernible P waves on the ECG.

**Aims:** To develop and prognostically validate a revised, explainable A-ECG heart age gap without incorporating P-wave measures.

**Methods:** An A-ECG heart age without P-wave measures (non-P) was derived from the 10-second 12-lead ECG in a derivation cohort using multivariable regression using an existing Bayesian 5-minute 12-lead A-ECG heart age as reference. The non-P heart age was externally validated in a separate cohort of patients referred for cardiovascular magnetic resonance imaging by describing its association with heart failure hospitalization or death using Cox regression, and its association with comorbidities.

**Results:** In the derivation cohort (n=2771), A-ECG non-P heart age agreed with the 5-min heart age (R^2^=0.91, bias 0.0±6.7 years), and increased with increasing co-morbidity. In the validation cohort (n=731, mean age 54±15 years, 43% female, n=139 events over 5.7 [4.8–6.7] years follow-up), increased A-ECG non-P heart age gap (≥10 years) associated with events (hazard ratio [95% confidence interval] 2.04 [1.38–3.00], C-statistic 0.58 [0.54–0.62], and the presence of hypertension, diabetes mellitus, hypercholesterolemia, and heart failure (p≤0.009 for all).

**Conclusions:** An explainable A-ECG non-P heart age gap applicable to both sinus and non-sinus rhythm associates with cardiovascular risk, cardiovascular morbidity, and survival.

## Introduction

Primary prevention in cardiovascular disease has underpinned the reduction in cardiovascular death over the decades and stands out as one of the most effective strategies for reducing its burden (1–3). Guidelines generally encourage both systematic and opportunistic cardiovascular screening with a focus on high-risk individuals (2, 4, 5). Despite being the simplest and most accessible cardiovascular diagnostic modality, current guidelines advise against the use of the traditional 12-lead electrocardiogram (ECG) in population screening due to its limited diagnostic performance (6). However, the ECG is known to contain information that is neither visually apparent nor immediately extractable with strictly conventional ECG analysis methods (7). Advanced ECG (A-ECG) analysis is one method of extracting such information from the ECG that offers markedly improved diagnostic performance over conventional ECG analysis, and could potentially enable more effective presymptomatic screening (8).

A-ECG analysis using high-fidelity 5-minute ECG recordings has been used to develop an accurate estimation of heart age concordant with the chronological age in healthy volunteers. ECG heart age by this method incrementally deviates from chronological age in individuals with cardiovascular risk factors and those with established cardiovascular disease (9). Subsequently, A-ECG analysis of the standard 10-second ECG has been used to derive an A-ECG heart age gap defined as the difference between A-ECG heart age and chronological age (10, 11). Increasing heart age gap has been shown to be associated with cardiovascular risk factors, heart failure and mortality (11). However, the presented A-ECG heart age estimation relied on presence of quantifiable P waves, limiting its utility to individuals in sinus rhythm. We hypothesized that heart age could also be derived using a standard 10-s ECG without including P-wave information. We therefore aimed to evaluate whether 10-s A-ECG could reliably predict heart age estimated from 5-min A-ECG, while not using any information from the P wave. We also aimed to externally validate the non-P-wave heart age gap estimation by determining the association with conventional risk factors and heart failure hospitalization or death.

## Methods

For this study, two pre-existing databases were used. For the derivation of heart age based on 10-second resting 12-lead ECG information excluding P-wave information, a database consisting of ECGs from 2,771 subjects (1,682 healthy volunteers, 305 individuals with cardiovascular risk factors, and 784 patients with established cardiovascular disease) was used (8, 9). The healthy subjects were recruited at Johnson Space Center (USA), the Universidad de los Andes (Venezuela), the University of Ljubljana hospitals and clinics (Slovenia), or Lund University Hospital (Sweden). Individuals included in the healthy cohort were low risk, asymptomatic volunteers with no identifiable cardiovascular or systemic disease based on clinical history and physical examination. Furthermore, active smokers, those with increased blood pressure at physical examination (≥140/90 mm Hg), and those on treatment for hypertension or diabetes were excluded from the healthy cohort.

Within the derivation dataset, Patients with cardiovascular risk factors (such as hypertension, diabetes mellitus, hypercholesterolemia or obesity) or established cardiovascular disease were recruited from cardiology clinics at either Texas Heart Institute (Houston, USA); the University of Texas Medical Branch (Galveston, USA), the University of Texas Health Sciences Centre (San Antonio, USA), Brooke Army Medical Centre (San Antonio, USA); St. Francis Hospital (Charleston, USA), the Universidad de los Andes (Mérida, Venezuela); and Lund University Hospital (Lund, Sweden). Inclusion in the established cardiovascular disease cohort was based on the presence of any of the following: 1) coronary heart disease, determined by coronary angiography with at least one obstructed vessel (≥ 50%) in at least one major coronary vessel, history of coronary artery bypass, or alternatively one or more reversible perfusion defects on ^99m^Tc-tetrofosmin single-photon emission computed tomography (SPECT); 2) left ventricular hypertrophy (LVH) based on imaging evidence of at least moderate, concentric wall thickening according to guidelines of the American Society of Echocardiography; 3) left ventricular systolic dysfunction (left ventricular ejection fraction ≤ 50%) at echocardiography, cardiac magnetic resonance imaging (CMR) or SPECT, with findings suggestive of ischemic or non-ischemic cardiomyopathy; or 4) echocardiographic or CMR-related findings consistent with hypertrophic cardiomyopathy. Details of the derivation cohort have been previously published (8). All patients in the derivation cohort underwent both 5-min and 10-s A-ECG analysis.

A second cohort was used for the purpose of assessing the prognostic value of the A-ECG non-P heart age gap produced from the derivation cohort. This validation cohort consisted of ECGs from patients who had undergone clinical CMR imaging at University of Pittsburgh Medical Centre (UPMC, Pittsburgh, PA, USA). All patients had 12-lead 10-sECG recorded within 30 days of the CMR exam and follow-up for death or hospitalization for heart failure. In the validation cohort, patients with missing follow-up data, heart rate ≥100/min, QRS duration ≥130 ms, atrial fibrillation or flutter, or digoxin use were excluded from the analysis. The validation cohort has been described in previous studies (12, 13). For the derivation cohort, Institutional Review Board (IRB) approvals were obtained from NASA’s Johnson Space Centre and partner hospitals that fall under IRB exemptions for previously collected and de-identified data. For the validation cohort, approval was obtained from the IRB at University of Pittsburgh Medical Centre. Written consent was obtained for all participants for both cohorts, and all data was analysed following de-identification.

### Heart Age Estimation in the Derivation Cohort

Both 5-minute and 10-second A-ECG analyses were applied to all ECGs in the derivation cohort. A-ECG measures obtained can be grouped into three categories: 1) Conventional ECG measures, such as heart rate, waveform durations, as well as frontal plane QRS- and T-wave axes, and amplitude-based criteria such as the Cornell or Sokolow-Lyon indices; 2) Spatial information from transformation of the 12-lead ECG to a derived vectorcardiogram using Kors’ transformation, including the spatial mean and maximum QRS-T angles, spatial azimuths and elevations, and the spatial ventricular gradient and its components, and 3) Waveform complexity measures of the QRS and T waves derived by singular value decomposition. A 10-s 12-lead ECG heart age has been described previously(10). For the purpose of this study, 10-second heart age was again derived using the 5-min Bayesian A-ECG heart age as the reference standard, but this time without allowing any P-wave information into the 10-second ECG approximation of the 5-minute score. The finally determined “non-P” 10-s A-ECG heart age score (formula) was then applied forward to all patients in the validation dataset.

### Statistical Analysis

Continuous variables were described using either mean±SD or median [interquartile range]. The chi-squared test was used to test for proportional differences between groups. When deriving the 10-s A-ECG “non-P” heart-age score, variable selection was determined in the healthy cohort through stepwise standard least squares multivariable linear regression for the prediction of the Bayesian 5-min heart age as the reference standard. Selection of variables was based on achieving the most parsimonious multivariable model with the highest possible model R^2^ with individually statistically significant (p<0.0001) measures. The intercept and coefficients of the prediction model was subsequently determined after applying the final model to both healthy subjects, patients with cardiovascular risk factors, and established cardiovascular disease using the 5-minute Bayesian A-ECG Heart Age as the reference standard.

Time-to-event analysis was conducted in the validation cohort, and Kaplan-Meier curves were constructed with censoring at the study’s end. The subjects were divided into two categories based on their non-P heart age gap: those with a gap of less than 10 years and those with a gap of 10 years or more, where 10 years approximately represents two standard deviations from the average heart age gap among healthy subjects in the initial derivation cohort (10). The association between the heart age gap and a composite endpoint of hospitalization for heart failure or death was analysed using Cox proportional hazard regression, unadjusted and adjusted for age, sex and cardiovascular risk factors (smoking, diabetes mellitus, hypertension, hypercholesterolemia and body mass index). A potential interaction effect between age and heart age gap on the association with outcomes was evaluated by analysing models with and without an interaction term for age (heart age gap × age). These models were then compared using the likelihood ratio test. Hazard ratios (HRs) are presented with 95% confidence intervals (CIs) for each 5-year increment in the heart age gap. A restricted cubic spline regression model was used to demonstrate risk increments with increasing heart age gap. Repeatability was assessed by describing the minimum detectable change (1.96 × √2 × standard error of the mean difference) in heart age gap one year later in a subset of healthy patients (n=27) for both the P-wave inclusive and P-wave exclusive heart age estimation. These patients all remained healthy at the time of the repeat ECG recording(14). These results are presented in Supplements, Table S1. Regression analysis and the derivation of the A-ECG Heart Age model was performed using SAS JMP version 11.0 (SAS Institute Inc, Cary, NC, USA). All other analyses were performed using the software R (version 4.2.2, R Core Team, R Foundation for Statistical Computing, Vienna, Austria).

## Results

### Derivation of the Non-P Heart Age

Characteristics of the derivation cohort are summarized in Table 1. The final non-P A-ECG heart age model with its ten (10) included measures are presented with intercepts and coefficients in Tables 2 and 3 for males and females, respectively. The mean values for the included A-ECG measures are summarized in Table 4, stratified by state of health, risk factors, and disease. The agreement between the non-P heart age and the 5-min heart age reference standard was strong (R^2^=0.91, bias 0.0±6.7 years, Figure 1). In healthy individuals, non-P gap was 0.4±5.0 years. in individuals with risk factors it was 7.4±6.8 years, and among those with established cardiovascular disease it was 13.5±8.3 years (Figure 2).

**Table 1.** Derivation Cohort Characteristics.

|  | All<br>(n = 2771) | Healthy<br>(n = 1682) | CV risk factors<br>(n = 305) | CV disease<br>(n = 784) |
| --- | --- | --- | --- | --- |
| Age, years | 46.2±16.0 | 38.6±13.0 | 54.8±11.2 | 59.0±13.2 |
| Male sex, n (%) | 1645 (59.4) | 999 (59.4) | 163 (53.4) | 483 (61.6) |
| Heart Age Gap, years | 4.9±8.6 | 0.4±5.0 | 7.4±6.8 | 13.5±8.3 |
| CAD, n (%) | 421 (15.2) | — | — | 421 (53.7) |
| ICM, n (%) | 120 (94.3) | — | — | 120 (15.3) |
| HCM, n (%) | 92 (3.3) | — | — | 92 (11.7) |
| LVH, n (%) | 96 (3.5) | — | — | 96 (12.2) |
| NICM, n (%) | 53 (1.9) | — | — | 53 (6.8) |

CAD: coronary artery disease; ICM: ischemic cardiomyopathy; HCM: hypertrophic cardiomyopathy; LVH: left ventricular hypertrophy; NICM: non-ischemic cardiomyopathy.

**Table 2.**
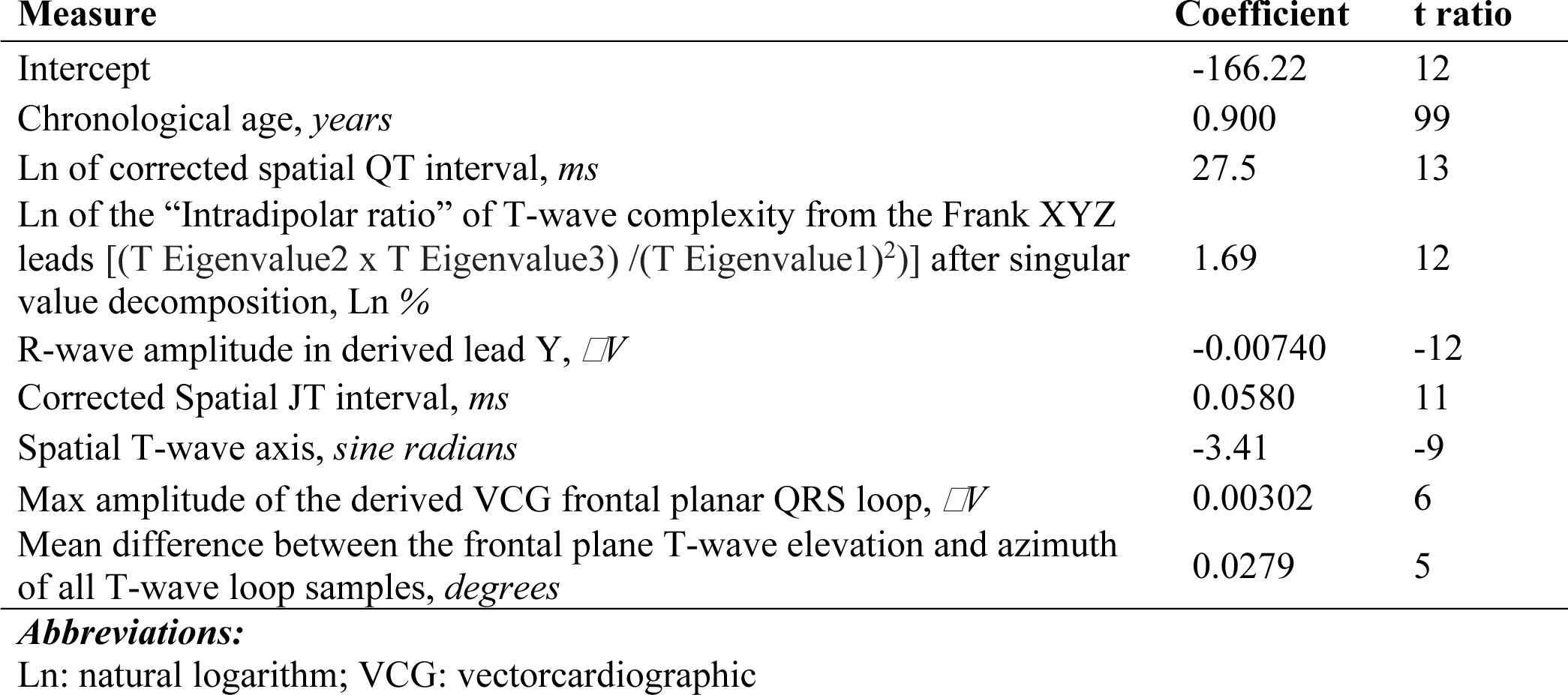
A-ECG measures included i n the non-P heart age for males.

| Measure | Coefficient | t ratio |
| --- | --- | --- |
| Intercept | -166.22 | 12 |
| Chronological age, *years* | 0.900 | 99 |
| Ln of corrected spatial QT interval, *ms* | 27.5 | 13 |
| Ln of the “Intradipolar ratio” of T-wave complexity from the Frank XYZ leads [(T Eigenvalue2 x T Eigenvalue3) / (T Eigenvalue1)<sup>2</sup>] after singular value decomposition, Ln % | 1.69 | 12 |
| R-wave amplitude in derived lead Y, $\mu V$ | -0.00740 | -12 |
| Corrected Spatial JT interval, *ms* | 0.0580 | 11 |
| Spatial T-wave axis, *sine radians* | -3.41 | -9 |
| Max amplitude of the derived VCG frontal planar QRS loop, $\mu V$ | 0.00302 | 6 |
| Mean difference between the frontal plane T-wave elevation and azimuth of all T-wave loop samples, *degrees* | 0.0279 | 5 |

**Abbreviations:**
Ln: natural logarithm; VCG: vectorcardiographic

**Table 3.** A-ECG measures included i n the non-P heart age for females.

| **Measure** | **Coefficient** | **t ratio** |
| --- | --- | --- |
| Intercept | -200 |  |
| Chronological Age, *years* | 0.942 | 91 |
| VCG Frontal plane QRS axis, *sine radians* | -9.59 | -23 |
| Gender | 4.20 | 14 |
| Natural logarithm Ln of corrected spatial QT interval, *ms* | 32.3 | 13 |
| R-wave amplitude in lead Y, $\mu V$ | -0.00869 | -12 |
| Ln of the “Intradipolar ratio” of T-wave complexity from the Frank XYZ leads $[(T \text{ Eigenvalue2} \times T \text{ Eigenvalue3}) / (T \text{ Eigenvalue1})^2]$ after singular value decomposition, Ln % | 1.88 | 12 |
| Corrected Spatial JT interval, *ms* | 0.0676 | 11 |
| Spatial T-wave axis, *sine radians* | -3.77 | -9 |
| Max amplitude of the derived VCG frontal planar QRS loop, $\mu V$ | 0.00 345 | 6 |
| Mean difference between the frontal plane T-wave elevation and azimuth of all T-wave loop samples, *degrees* | 0.0334 | 5 |

**Abbreviations:**
Ln: natural logarithm; VCG: vectorcardiographic

**Table 4.** Advanced ECG parameters in the derivation cohort stratified by health and presence of cardiovascular risk factors or established cardiovascular disease.

|  | Healthy | Risk<br>Factors | Disease | *p* |
| --- | --- | --- | --- | --- |
| n (%) | 1,682 (60.7) | 305 (11) | 784 (28.3) |  |
| Chronological age, *years* | 39±13 | 55±11 | 59±13 | <0.001 |
| R-wave amplitude in lead Y, $\mu V$ | 1144±392 | 828±363 | 717±462 | <0.001 |
| VCG frontal plane QRS axis, *sine radians* | 0.82±0.23 | 0.54±0.44 | 0.29±0.54 | <0.001 |
| Max amplitude of the derived VCG frontal planar QRS loop, $\mu V$ | 1617±411 | 1384±385 | 1286±599 | <0.001 |
| Ln of the “Intradipolar ratio” of T-wave complexity from the Frank XYZ leads [(T Eigenvalue2 x T Eigenvalue3) / (T Eigenvalue1)<sup>2</sup>] after singular value decomposition, Ln % | -0.69±0.83 | -0.28±0.89 | 0.43±1.12 | <0.001 |
| Ln of the corrected spatial QT interval, *ms* | 6.00 (0.06) | 6.05 (0.06) | 6.07 (0.08) | <0.001 |
| Spatial T-wave axis, *sine radians* | 0.71 (0.20) | 0.63 (0.28) | 0.46 (0.54) | <0.001 |
| Corrected spatial JT interval, *ms* | 336±23 | 355±29 | 364±37 | <0.001 |
| Mean difference between the frontal plane T-wave elevation and azimuth of all T-wave loop samples, *degrees* | 41±16 | 47±22 | 72±36 | <0.001 |

**Abbreviations:**
Ln: natural logarithm; VCG: vectorcardiographic

**Figure 1.**
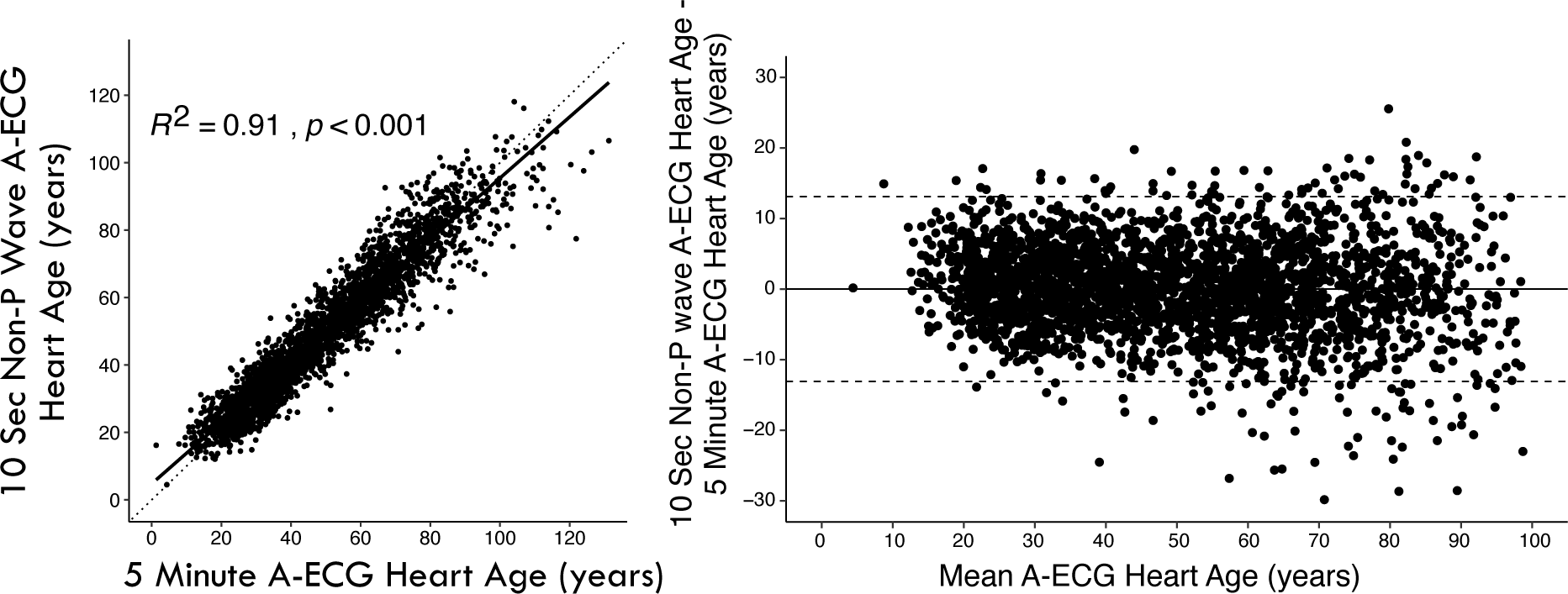
Left panel: Scatter plot showing the relationship between the 10-s, Non-P-wave Advanced-ECG (A-ECG) heart age and the 5-min A-ECG heart age in the derivation cohort. The R^2^ value was 0.91 (p < 0.001). Right panel: Bland–Altman plot showing the difference between the 10-s Non-P wave A-ECG Heart Age and 5-min A-ECG heart age in relation to the mean of both ECG heart ages. The agreement between methods is strong, with minimal deviation from the identity line (dashed) or bias (0.0 ± 6.7 years).

**Figure 2.**
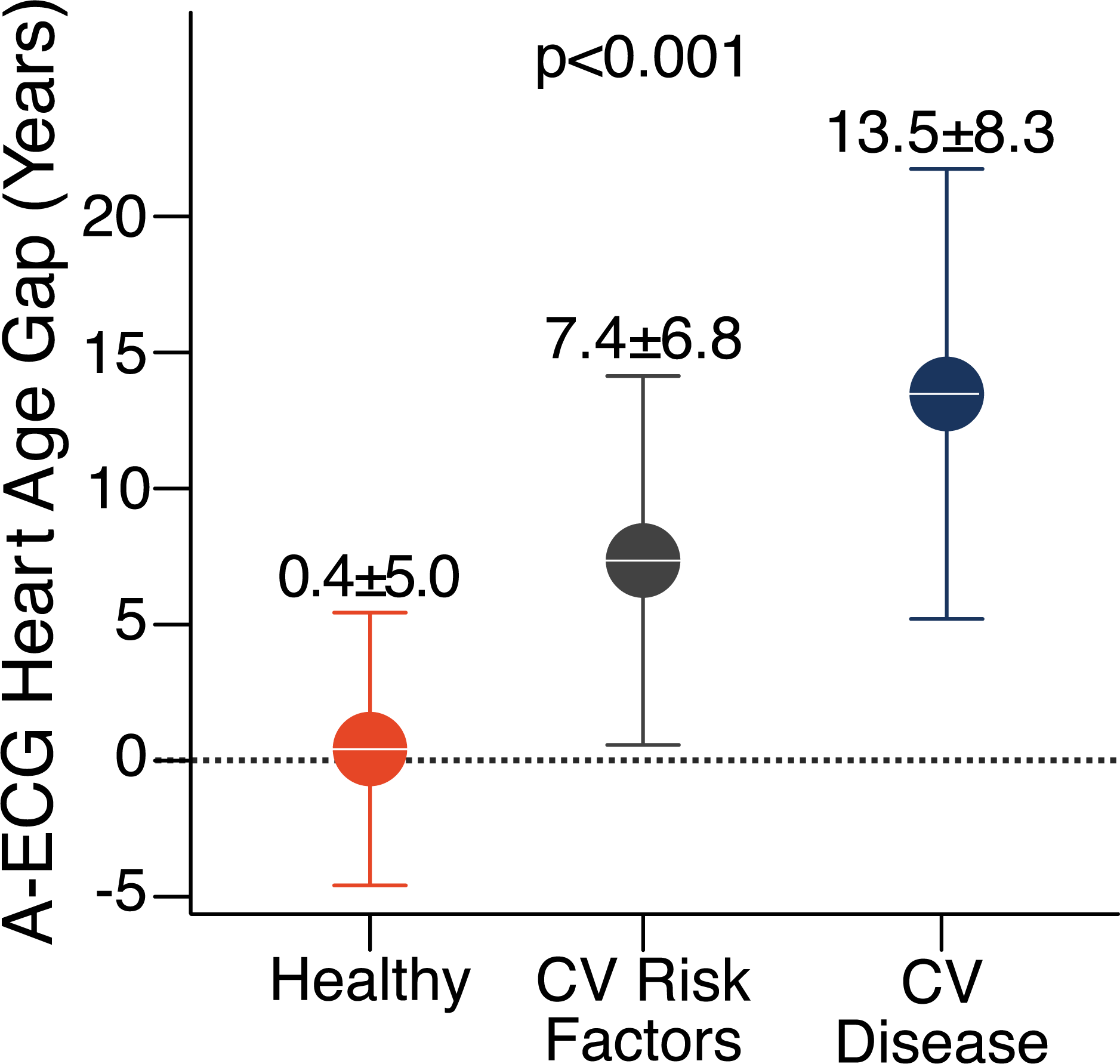
Advanced ECG (A-ECG) non-P heart age gap in healthy individuals (left, orange), individuals with cardiovascular (CV) risk factors (middle, dark grey), and patients with CV disease (right, navy blue). On average, there is a negligible difference between non-P heart age and chronological age in healthy individuals, whereas the gap is increased in individuals at CV risk and highest for those with overt CV disease.

**Figure 3.**
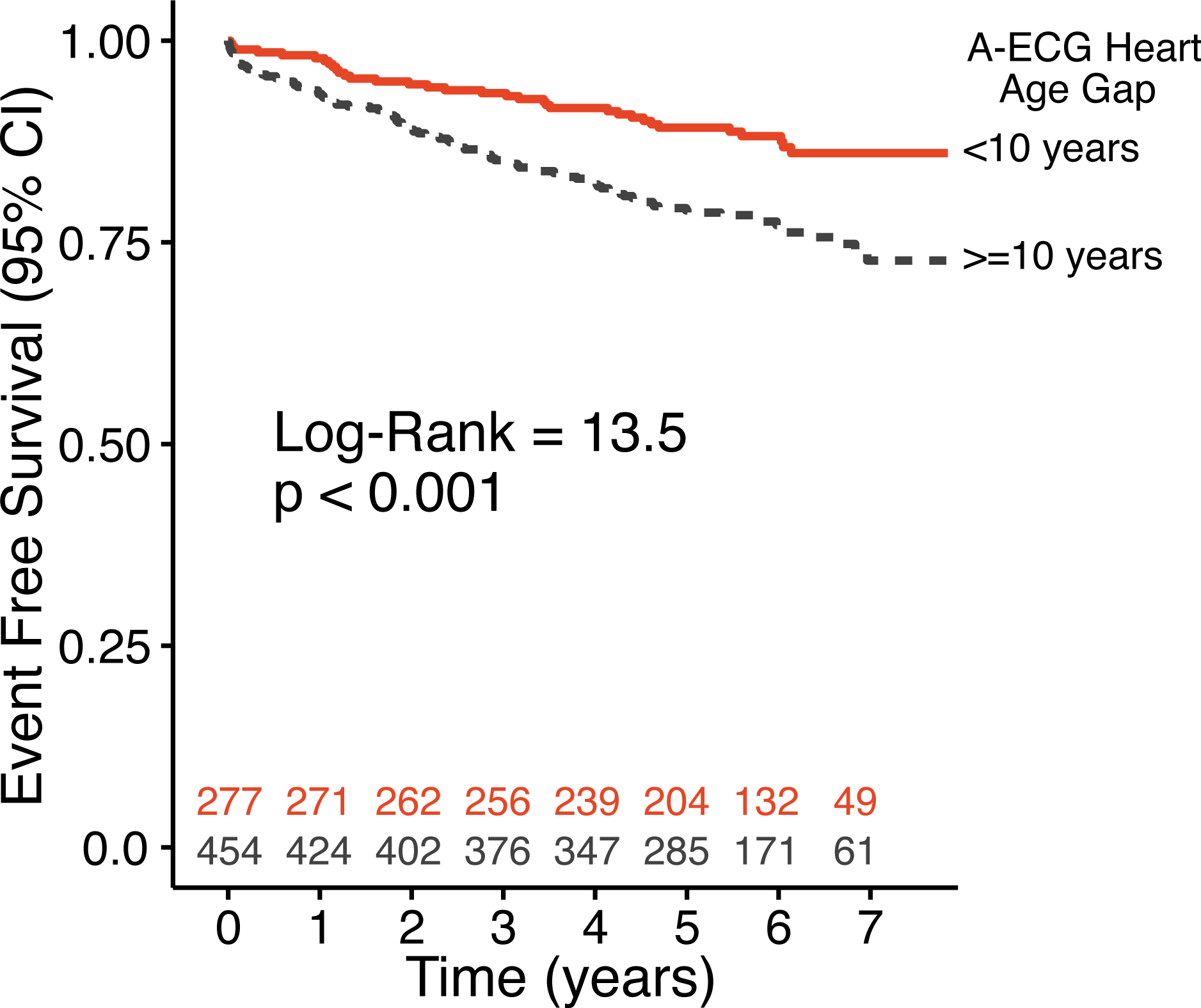
Time-to-event analysis for individuals with A-ECG non-P heart age gap<10 years (dense, Orange line) versus those with an A-ECG non-P heart age gap>=10 years (dashed grey line) for hospitalization for heart failure or death in the external validation cohort.

**Figure 4.**
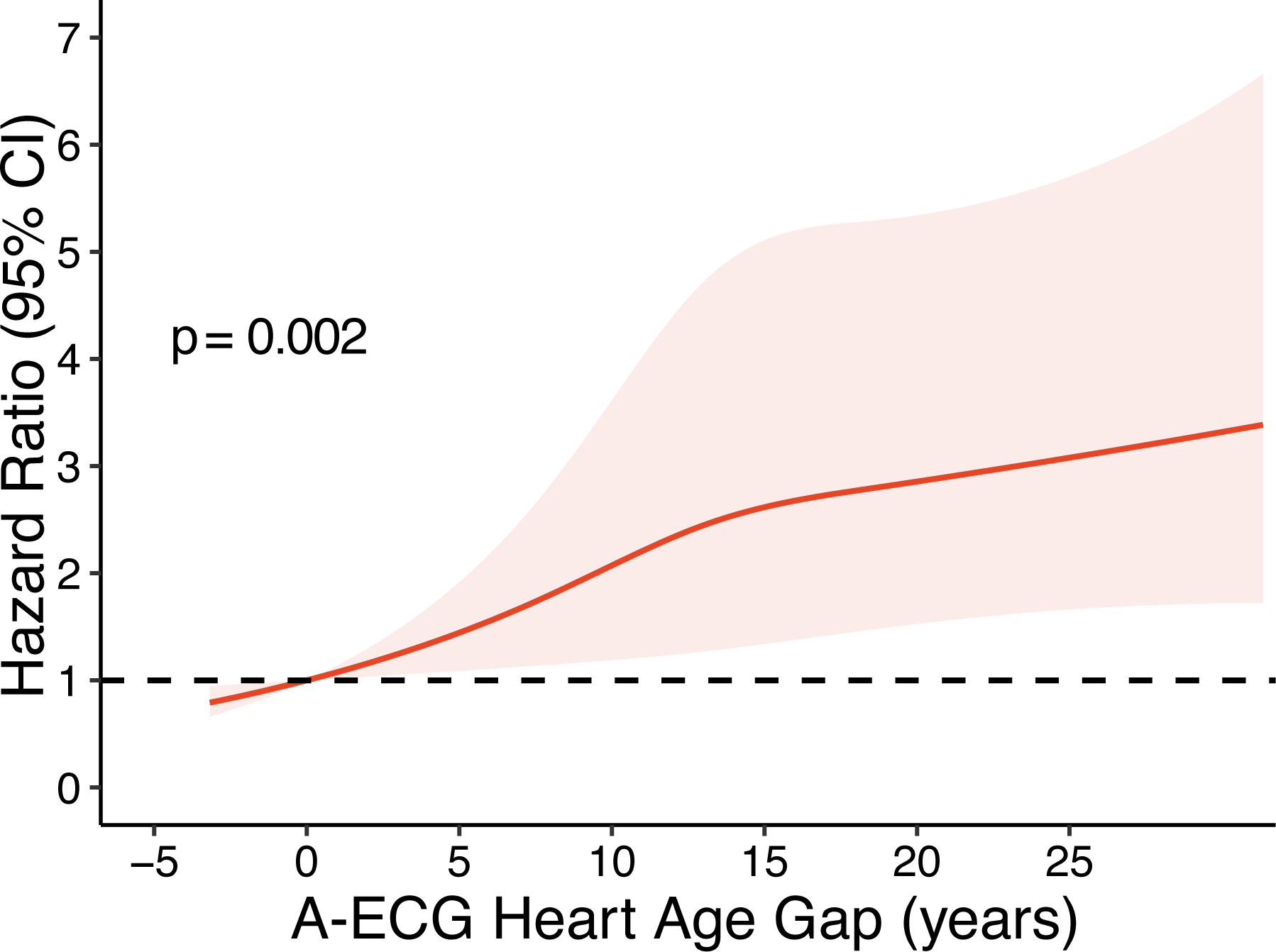
Restricted cubic splines plot showing the continuously increasing hazard ratio for death or hospitalization for heart failure with increasing non-P A-ECG heart age gap. The shaded area indicates the 95% confidence interval.

### External validation of the Non-P Heart Age Gap

The validation cohort included 731 patients with a median follow-up of 5.7 [4.8–6.7] years, during which 138 (18.9%) individuals experienced the primary combined outcome of hospitalization for heart failure or death (101(13.8%) deaths, 57(7.8%) hospitalizations for heart failure, and 20 (2.7%) both hospitalisation and death).Baseline characteristics of the validation cohort are presented in Table 5.

**Table 5.** Validation Cohort Detailed Characteristics.

|  | All Cohort | Non-P Heart Age Gap (Years) |  | p |
| --- | --- | --- | --- | --- |
|  |  | <10 | ≥10 |  |
| Number of individuals, n (%) | 731 (100) | 277 (37.9) | 454 (62.1) |  |
| Chronological Age, years | 53.6±15.3 | 49.6±15.2 | 56.0±14.8 | <0.001 |
| Non-P A-ECG Heart Age, years | 66.7±19 | 54.4±16.1 | 74.1±16.7 | <0.001 |
| Non-P A-ECG Heart Age Gap, years | 13.1±8.2 | 4.8±3.8 | 18.1±5.7 | <0.001 |
| Male Sex, n (%) | 417 (57.0) | 160 (57.8) | 257 (56.6) | 0.82 |
| Body-mass index, kg/m<sup>2</sup> | 30±7.8 | 29.4±7.6 | 30.3±7.8 | 0.12 |
| Body Surface Area, m<sup>2</sup> | 2.0±0.3 | 2.0±0.3 | 2.1±0.3 | 0.29 |
| All cause mortality, n (%) | 101 (13.8) | 28 (10.1) | 73 (16.1) | 0.031 |
| Death or HF Hospitalization, n (%) | 138 (18.9) | 34 (12.3) | 104 (22.9) | 0.001 |
| NT-proBNP, ng/L | 64 [28–188] | 43 [20–131] | 82 [33–222] | <0.001 |
| eGFR, mL/min/1.73m<sup>2</sup> | 89±25 | 93±23 | 86±26 | 0.001 |
| Hypertension, n (%) | 379 (51.8) | 120 (43.3) | 259 (57.0) | <0.001 |
| Diabetes Mellitus, n (%) | 157 (21.5) | 33 (11.9) | 124 (27.3) | <0.001 |
| Hyperlipidemia, n (%) | 295 (40.4) | 86 (31.0) | 209 (46.0) | <0.001 |
| Chronic Kidney Disease, n (%) | 18 (2.5) | 2 (0.7) | 16 (3.5) | 0.034 |
| Smoker, n (%) | 119 (16.3) | 50 (18.1) | 69 (15.2) | 0.36 |
| CABG, n (%) | 56 (7.7) | 20 (7.2) | 36 (7.9) | 0.84 |
| PCI, n (%) | 89 (12.2) | 25 (9.0) | 64 (14.1) | 0.06 |
| Heart Failure Hospitalizations, n (%) | 57 (7.8) | 11 (4.0) | 46 (10.1) | 0.004 |
| HFrEF, n (%) | 166 (22.7) | 48 (17.3) | 118 (26.0) | 0.009 |
| HFpEF, n (%) | 51 (7) | 13 (4.7) | 38 (8.4) | 0.081 |
| *Cardiovascular Magnetic Resonance Imaging Parameters* |  |  |  |  |
| End-diastolic LV volume, mL | 172±63 | 167±57 | 175±67 | 0.084 |
| End-systolic LV volume, mL | 83±58 | 75±47 | 85±63 | 0.003 |
| LVEF, % | 55±14 | 57±12 | 54±15 | 0.002 |
| GLS, % | -16±4 | -16±4 | -15±5 | <0.001 |
| LV mass, g | 120±46 | 115±42 | 124±49 | 0.009 |
| Presence of LGE, % | 263 (36) | 83 (30.0) | 180 (39.6) | 0.010 |
| Extracellular volume, % | 27.9 (3.8) | 27.4 (3.5) | 28.2 (4) | 0.008 |
| *Medications* |  |  |  |  |
| Insulin, n (%) | 107 (14.6) | 23 (8.3) | 84 (18.5) | <0.001 |
| Oral Hypoglycemic, n (%) | 49 (6.7) | 12 (4.3) | 37 (8.1) | 0.06 |
| ACE/ARB, n (%) | 295 (40.4) | 110 (39.7) | 185 (40.7) | 0.84 |
| No Medications, n (%) | 133 (18.2) | 73 (26.4) | 60 (13.2) | <0.001 |
| Beta Blockers, n (%) | 359 (49.1) | 109 (39.4) | 250 (55.1) | <0.001 |
| Loop Diuretics, n (%) | 142 (19.4) | 39 (14.1) | 103 (22.7) | 0.006 |
| Calcium Channel Blocker, n (%) | 55 ( 7.5) | 14 (5.1) | 41 (9.0) | 0.067 |
| Hydrochlorothiazide, n (%) | 68 ( 9.3) | 22 (7.9) | 46 (10.1) | 0.39 |
| Nitroglycerin, n (%) | 23 ( 3.1) | 6 (2.2) | 17 (3.7) | 0.33 |
| Statin, n (%) | 301 (41.2) | 91 (32.9) | 210 (46.3) | <0.001 |
| Warfarin, n (%) | 45 ( 6.2) | 14 (5.1) | 31 (6.8) | 0.42 |
| Antiplatelets, n (%) | 379 (51.8) | 120 (43.3) | 259 (57.0) | <0.001 |

NT-proBNP: N-terminal prohormone of brain natriuretic peptide; eGFR: estimated glomerular filtration rate; CAD: coronary artery disease; CABG: coronary artery bypass grafting; PCI: percutaneous coronary intervention; HFrEF: heart failure with reduced ejection fraction; HFpEF: heart failure with preserved ejection fraction; GLS: global longitudinal strain; LGE: late gadolinium enhancement; LV: left ventricle; LVEF: left ventricular ejection fraction; ACE: angiotensin-converting enzyme; ARB: Angiotensin Receptor Blockers;.

Cardiovascular risk factors and diseases were more prevalent among patients with a higher gap (≥10 years) compared to those with a lower gap (<10 years; hypertension 57%vs 43%, p<0.001; diabetes 27%vs 12% p<0.001; hyperlipidaemia 46% vs 31%, p<0.001). In addition, abnormal CMR imaging findings were more prevalent among those with higher gap, compared to those with a lower gap (lower LVEF, worse global longitudinal strain (GLS), higher left ventricular mass, greater presence of late gadolinium enhancement, and higher myocardial extracellular volume, p<0.01 for all).

Increased gap (≥10 years) was significantly associated with increased risk of heart failure admission or death (unadjusted HR: 2.04 [1.38–3.00], adjusted for age and sex: 1.55 [1.04– 2.30]). The interaction test for gap and age did not achieve statistical significance (p=0.06). Given the close proximity to the predefined significance level (p<0.05), HRs were also calculated after stratifying data by age. Heart age Gap ≥10 years was associated with heart failure or death among younger individuals (<60 years: HR 2.98 [1.68–5.30]), but not among ≥60 years (HR 1.01 [0.60–1.71]).

## Discussion

The main finding of this study is that an estimation of ECG heart age containing useful prognostic value can be performed without the use of P-wave information. Healthy volunteers had a “non-P” heart age similar to their chronological age, i.e. their gap was close to zero. In comparison, individuals with cardiovascular risk factors had a gap of approximately 7 years, while those with established disease had a gap of 14 years. In the external validation cohort, high gap was associated with higher prevalence of cardiovascular risk factors, abnormal CMR findings, and ultimately hospitalization for heart failure or death.

The current study builds on prior work enabling the use of an A-ECG heart age on standard 10-second, 12-lead ECGs rather than a 5-minute, 12-lead higher-fidelity ECG as originally devised (9–11). However, P-wave duration constituted an important part of the initial 10-second A-ECG model for heart age, limiting its use to individuals in sinus rhythm (10). The current study therefore now extends the applicability of 10-second A-ECG heart age estimations to patients with atrial fibrillation or other causes of non-quantifiable P-waves.

The current study mechanistically shows that determinants of the emerging concept of cardiovascular ageing are embedded in the ECG along domains beyond the P wave, which has previously been described as an important measure associated with cardiovascular ageing and associated pathologies (15–18). In the original 10-second A-ECG heart age gap, P-wave and spatial QT durations were the two most important A-ECG measures in the model. By contrast, in the non-P heart age gap of the current study, the axis of the spatial QRS axis in the derived vectorcardiographic frontal plane and the heart rate-corrected spatial QT interval were the two most important ECG measures in the model. Indeed, the normal evolution of these two metrics with ageing is well known (19, 20).

Higher non-P gap was also associated with both adverse outcomes and underlying comorbidities. This is consistent with multiple recent studies on the concept of biological age estimation using the ECG (21–28).For example, a deep neural network (DNN) has been trained to predict sex and age from the 12-lead ECG. In that study, those with a predicted heart age >7 years higher than their chronological age (gap ≥7 years)had a higher prevalence of coronary disease, hypertension, and reduced left ventricular ejection fraction, while those with a lower gap had fewer events in long-term follow-up(21). Similarly, DNN-estimated ECG age trained on a large cohort (n=1,558,415) showed increased risk with increasing gap (27).

A-ECG emphasizes the use of transparently explainable and quantifiable ECG measures for heart age estimation. This differs from other attempts at heart age estimation using the ECG, particularly from DNN-type modelling. For example, DNN techniques continue to have the inherent limitation of model opaqueness and non-explainability and as such are still considered “black box” methods. In the only head-to-head comparison on an identical external validation cohort, the original 10-second A-ECG heart age outperformed a DNN-based heart age estimation model in prognostic strength (11). In terms of explainability, measures included in the non-P gap are transparently quantifiable digital biomarkers of cardiac electrophysiology. Some of these digital biomarkers have been described extensively in the literature, such as the frontal plane QRS axis, spatial QT duration, vectorcardiographic T-wave axis, spatial ventricular gradient and certain ratios between the T-wave eigenvalues that quantify the complexity of the T wave in three-dimensional space. Furthermore, the deviations of these measures from normal ranges are known to be associated with adverse events (29–35).

In this heart age model, chronological age was included and used as the foundation from which accelerated or decelerated cardiac aging then occurred based on electrocardiographic deviations from healthy aging. In contrast, DNN-type heart age models have focused on predicting the actual chronological age of study participants (11).

The non-p heart age was derived using strictly healthy individuals in whom cardiovascular risk factors or disease were actively ruled out. Hence, no disease signatures were embedded in the ECGs used for training the model. Beyond scientific curiosity, there appears to be little clinical relevance in predicting the actual chronological age from an ECG among patients with known or suspected cardiovascular disease. Interestingly, the association between non-P gap and the primary outcome was stronger in younger individuals who should otherwise be at lower risk of adverse cardiovascular events. In younger individuals, the association between heart age gap and prognosis is consistent with this association being attributable to risk factors and disease. In contrast, among older individuals, it is possible that a higher prevalence of other risk factors or established disease lessen the prognostic impact of the heart age gap.

The findings of the current study align with multiple previous publications on electrocardiographic signals of accelerated ageing. The emerging understanding from DNA methylation studies of the process of accelerated cellular senescence in response to adverse lifestyle and external factors indicate that perhaps the ECG can be an accessible and intuitive ‘epigenetic clock’ with clinical and public health utility (35–38). This theory is further supported by findings of a weak albeit linear relationship between accelerated ageing as measured using DNA methylation-based ageing biomarkers and A-ECG Heart Age (40). A higher ECG heart age gap has been shown to be associated with Lamin-related gene (LMNA gene) mutations related to accelerated aging in Progeria syndrome (39), as well as impaired peripheral microvascular endothelial dysfunction, which is associated with vascular ageing (22). Based on these observations, the association between the non-P gap and cardiovascular morbidity and mortality possibly stems from detecting subtle electrophysiological signatures related to accelerated cellular senescence.

The greater purpose of non-P gap is to have easy access to a simple yet prognostic and explainable biomarker of cardiovascular risk that can be easily deployed anywhere standard ECGs are performed, and intuitively communicated to a patient to aid personalized decision-making and intervention. For example, informing an individual with unaddressed risk factors that their heart is 15 years older than their chronological age could be a powerful message to spur preventive action. Similar attempts have been made in other areas, such as to incentivize smoking cessation through age-estimations based on pulmonary function testing results (42). Beyond the mechanistic insights offered by the non-P gap, it extends its use to a more diverse population, particularly those in non-sinus rhythm. Whether heart age estimations in patients in atrial fibrillation can improve thromboembolic risk stratification merits testing in future studies.

The current non-P gap is the first heart age gap specifically developed for applications in non-sinus rhythms. Atrial fibrillation is a widely heterogenous condition and also the most common non-sinus rhythm. Potential applications of the heart age gap in atrial fibrillation include phenotyping (43), risk stratification at time of diagnosis, and monitoring the effects of therapy, with future studies of these applications being justified.

### Limitations

The validation cohort consisted of patients referred for CMR imaging, representing a group with a higher prevalence of cardiovascular risk factors and diseases compared to an otherwise relatively asymptomatic general practice outpatient population who could be a target for clinical application of the non-P gap. Despite this, the non-P gap retained its simplicity and utility even in this higher-risk group. Additionally, the cardiovascular risk factors and conditions observed in this cohort are representative of the very issues that preventive healthcare measures strive to address.

Although the purpose of developing a heart age gap without P wave measures is to enable heart age estimation in patients with non-sinus rhythms, the validation was performed in patients in sinus rhythm. Therefore, the results of this study should only be considered as proof-of-concept at this stage regarding applicability in non-sinus rhythms. Further validation studies in individuals in non-sinus rhythms are justified.

### Conclusions

Accurate and prognostically useful heart age estimations can be made through A-ECG analysis applied to standard 10-second resting 12-lead ECGs without relying on P-wave-derived measures. This therefore now extends the applicability of A-ECG heart age estimations to patients who are in non-sinus rhythms.

## Data Availability

Deidentified data of this study can be made available upon reasonable request to the corresponding author.

---

**Supplemental Table 1.** Serial A-ECG non-P heart age and p-wave heart age for healthy individuals performed 1 year apart.

|  | **Non-p heart age** | **p-wave heart age** |
| --- | --- | --- |
| **Number of subjects, n** | 27 | 27 |
| **Males, n (%)** | 14 (52) | 14 (52) |
| **Age, years** | 40.5±9 | 40.5±9 |
| **Heart age gap at first measurement, years** | -1.4±4.5 | -1.6±4.0 |
| **Difference between heart age gap one year apart, years** | 0.4±1.8 | 0.6±1.9 |
| **Intraclass correlation coefficient, unitless** | 0.93 [0.84-0.97] | 0.88 [0.76–0.95] |
| **Minimum detectable change, years** | 0.9 [0.3-1.6] | 1.0 [0.3–1.7] |

## Notes

### Competing Interest Statement

The authors have declared no competing interest.

### Funding Statement

This study did not receive any funding.

### Author Declarations

For the derivation cohort, Institutional Review Board (IRB) approvals were obtained from NASA's Johnson Space Center and partner hospitals that fall under IRB exemptions for previously collected and de-identified data. For the validation cohort, approval was obtained from the IRB at University of Pittsburgh Medical Center. Written consent was obtained for all participants for both cohorts, and all data was analyzed following de-identification.

